# Cerebrospinal fluid dopa-decarboxylase is a biomarker of dopaminergic dysfunction

**DOI:** 10.64898/2026.10.13.26364985

**Authors:** Silvia Sperandei, Federico Paolini Paoletti, Megan De Pauw, Lorenzo Gaetani, Cristina Tranfaglia, Julie Goossens, Erik Stoops, Eugeen Vanmechelen, Lucilla Parnetti, Giovanni Bellomo, Davide Chiasserini

## Abstract

**Background:** Nigrostriatal dopaminergic dysfunction, a biological anchor of Parkinson’s disease (PD), is routinely assessed using single-photon emission computed tomography of the dopamine transporter (DaT-SPECT). Dopa decarboxylase (DDC) in cerebrospinal fluid (CSF) is a promising biomarker of dopaminergic dysfunction, but its association with DaT-SPECT has to be demonstrated.

**Objectives:** To investigate the association of CSF DDC with DaT-SPECT.

**Methods:** We first analysed proteomic and DaT-SPECT data from the Parkinson’s Progression Markers Initiative (PPMI), including PD (n=891), prodromal PD (n=867) and healthy controls (n=161). We then verified this association in a single-center clinical cohort (UNIPG), including PD/Dementia with Lewy Bodies (DLB) (n=78), atypical parkinsonisms (AP, n=11), other neurodegenerative diseases (OND, n=25) and controls (CTRL, n=40). Here, CSF DDC was measured by a novel fully automated quantitative immunoassay.

**Results:** In PPMI, among 129 proteins assessed, DDC showed the strongest association with DaT-SPECT (β = −0.132, FDR = 6.92 × 10^−33^). Across PPMI and UNIPG cohorts, DDC correlated with striatal binding ratio (SBR) (ρ = −0.377, P < 0.001; ρ = −0.466, P < 0.001, respectively) and accurately discriminated positive from negative scans (AUC 0.811, 95% CI 0.762–0.859; AUC 0.870, 95% CI 0.777–0.963, respectively). In PPMI prodromal cases, higher DDC predicted faster motor progression. In UNIPG, DDC distinguished PD/DLB and AP from OND and CTRL groups (AUC 0.830, 95% CI 0.764–0.896), independently of dopaminergic treatment.

**Conclusion:** CSF DDC reflects nigrostriatal dopaminergic dysfunction, representing a potential quantitative biochemical alternative to DaT-SPECT that might be readily implemented via automated immunoassays.

## Introduction

Nigrostriatal dopaminergic dysfunction is the pathogenetic hallmark of Parkinson’s Disease (PD). The nuclear imaging of the dopaminergic system is extensively used to objectify this feature for diagnostic purposes. Single-Photon Emission Computed Tomography (SPECT) imaging with ioflupane (I^123^) to visualize dopamine transporter (DaT) is currently the most widely used approach in clinical practice^1^.

According to the proposed biological staging framework for PD^2^, the two main anchors for diagnosing PD are represented by the presence of α-synuclein pathology (S+) - detected by means of α-synuclein seed amplification assays (α-syn-SAA) either in cerebrospinal fluid (CSF) or in skin biopsy - and the dopaminergic dysfunction (D+) due to nigrostriatal degeneration.

Within this framework, a quantitative DaT-SPECT measure - namely, the age- and sex-adjusted striatal binding ratio (SBR) in the most affected putamen - has been proposed as an indicator of dopaminergic deficit.

CSF DOPA-decarboxylase (DDC), the enzyme responsible for the synthesis of dopamine from L-DOPA^3^, might represent a biochemical alternative to DaT-SPECT, which is relatively costly, time-consuming and requires exposure to radioactive tracers and ionizing radiation. Available data already show that CSF DDC accurately discriminates parkinsonian syndromes from healthy controls and other neurodegenerative disorders^4,5^, also correlating with disease severity in PD and dementia with Lewy bodies (DLB)^5^. On the other hand, the evidence on the relationship between CSF DDC levels and dopaminergic dysfunction assessed by DaT-SPECT is still insufficient. Some studies reported poor association^5^ while, in other reports, the correlation was influenced by dopaminergic treatment^6,7^. The aim of the present study was to evaluate CSF DCC as a potential biochemical marker of dopaminergic dysfunction, alternative to DaT-SPECT. The investigation was conducted in two phases. We first investigated whether CSF DDC reflects striatal dopaminergic dysfunction using clinical, proteomic, and DaT-SPECT data from the Parkinson’s Progression Markers Initiative (PPMI), including patients with established PD, prodromal cases, and healthy controls. We then assessed this association in an independent real-world cohort from the University of Perugia (UNIPG), composed of patients with parkinsonian disorders and other neurological diseases. In this cohort CSF DDC was measured using a novel, fully automated quantitative immunoassay.

## Materials and methods

### PPMI cohort

PPMI data were obtained on June 15, 2026, from the Parkinson’s Progression Markers Initiative database (PPMI; http://www.ppmi-info.org/access-data-specimens/download-data; RRID: SCR_006431)^8^. Participants with available CSF proteomic data from the baseline study visit were included. CSF DDC was measured, together with 128 other proteins, using the CNS 120 NULISA® panel (Alamar Biosciences, Fremont, CA, USA) and expressed as normalized protein quantity (NPQ) values.

The NULISA proteomic dataset at baseline comprised 1,919 subjects: 891 patients with PD, 867 with prodromal PD and 161 CTRL. Quantitative DaT-SPECT striatal binding ratio (SBR) data were available for 1,606 participants and qualitative classification for 792 participants (Supplementary Table 1).

DaT-SPECT images were acquired according to a standardized protocol and centrally processed by the PPMI imaging core laboratory. Visual assessments classified scans as positive or negative for evidence of a presynaptic dopamine transporter deficit. Quantitative analyses included SBR values for the caudate nuclei, putamina and whole striata. ^9,10,11^ For each participant, the scan closest to CSF collection, within a maximum interval of 18 months, was included. Further technical details on dataset construction, clinical variables and imaging are provided in Supplementary Methods.

### UNIPG cohort

The UNIPG cohort included 114 consecutive patients with Parkinsonian syndromes or other neurodegenerative diseases (OND) who were referred to the Neurology Clinic of the University Hospital of Perugia for diagnostic purposes, with available CSF samples and DaT-SPECT acquired within 18 months from CSF collection. Lumbar puncture was carried out according to international guidelines^12^. Parkinsonian syndromes (n=89) comprised PD (n = 72), DLB (n = 6) and atypical parkinsonisms (AP, n = 11), including progressive supranuclear palsy (PSP, n=7) and multisystem atrophy (MSA, n=4). The OND group (n = 25) included patients with tremor and/or other motor symptoms whose final diagnosis, based on clinical and imaging findings, was not consistent with PD, DLB, or AP. The composition of the OND group is reported in **Supplementary table 2**. Six patients with an inconclusive DaT-SPECT visual read were excluded from qualitative analyses only; quantitative SBR data were retained.

For DDC measurements, we also included, as control group (CTRL), 40 subjects with isolated minor neurological conditions (i.e., mononeuropathy, headache and seizures), who underwent lumbar puncture as part of their routine diagnostic work-up.

DaT-SPECT scans were visually classified as positive or negative for evidence of presynaptic dopaminergic dysfunction. Quantitative analysis was performed using Basal Ganglia Matching Tools V2 (automated three-dimensional BasGan method), using occipital background reference and partial volume effect correction. Mean striatal SBR was calculated as the average of the bilateral caudate and putamen SBR^13^. Further clinical and imaging details are provided in the Supplementary Methods. All the procedures were performed in accordance with Declaration of Helsinki. All the patients and/or their legal representatives gave informed written consent for the lumbar puncture and the inclusion in the study that was approved by the local Ethics Committee (Comitato Etico Aziende Sanitarie Regione Umbria registry N°: 3944/21)

### A monoclonal antibody-based Lumipulse^®^ G DDC CSF immunoassay

In the UNIPG cohort, DDC concentrations in CSF were quantified using the Lumipulse® G ADxPLORER® DDC V1 assay (Fujirebio Europe N.V., Ghent, Belgium) on the LUMIPULSE® G1200 automated immunoassay platform. The assay was developed using the proprietary monoclonal antibody ADx412, which recognizes a C-terminal-specific epitope of DDC, as the capture antibody, and an alkaline phosphatase-conjugated Fab fragment derived from the ADx411 monoclonal antibody as the detector. The assay was calibrated with a recombinant DDC full-length protein, with a calibration range covering CSF DDC concentration from 1.5-600 pg/ml. Analytical qualification of this assay included assessment of sensitivity, precision, parallelism, and spike recovery. The lower functional limit of quantification (LLOQ) was 0.5 pg/mL, whereas the functional measurement range is 1.5-25 pg/mL. Mean intra- and inter-assay variation were 3.8% and 4.3%, respectively, using 5 samples (2 replicates over 5 runs). Parallelism and spike recovery met predefined acceptance criteria. UNIPG samples were analyzed in singlicate in a single run, blinded to clinical data. Further analytical details are provided in the Supplementary Methods.

### Statistical analysis

Statistical analyses used R 4.5.1 and Python 3.12. Data are summarized as median (IQR) or n (%). In PPMI, associations between CSF proteins and regional SBR were assessed using linear regression with z-standardized variables, adjusted for age, sex, and L-dopa Equivalent Daily Dose (LEDD), with Benjamini–Hochberg FDR correction applied separately to each imaging outcome. Diagnostic-group differences in DDC were assessed using age-adjusted linear models and FDR-corrected pairwise comparisons of estimated marginal means. Logistic regression modeled DaT-SPECT status as a function of DDC, age, and LEDD, excluding six equivocal UNIPG scans. ROC analyses estimated AUCs with DeLong 95% CIs and Youden-derived cut-offs. DDC–SBR associations were assessed using Spearman correlations with 95% CIs from 2,000 bootstrap resamples.

Longitudinal analyses included prodromal participants with isolated REM sleep behavior disorder or hyposmia, baseline Hoehn and Yahr stage 0, and MDS-UPDRS III <10. Progression was defined as the first MDS-UPDRS III score ≥10, with censoring at six years or the last available visit. Kaplan– Meier curves compared median-defined biomarker groups using log-rank tests. Cox models evaluated DDC and SBR as continuous standardized and median-dichotomized predictors, adjusting for age, sex, prodromal subgroup, and baseline MDS-UPDRS III, and reporting HRs with 95% CIs. All tests were two-sided, with significance set at p <0.05 using FDR-adjusted values where applicable.

## Results

### Demographic and clinical characteristics

Data on demographic features, clinical measures, LEDD and DaT-SPECT qualitative results for the PPMI and UNIPG cohorts are summarized in **Table 1**.

**Table 1.**
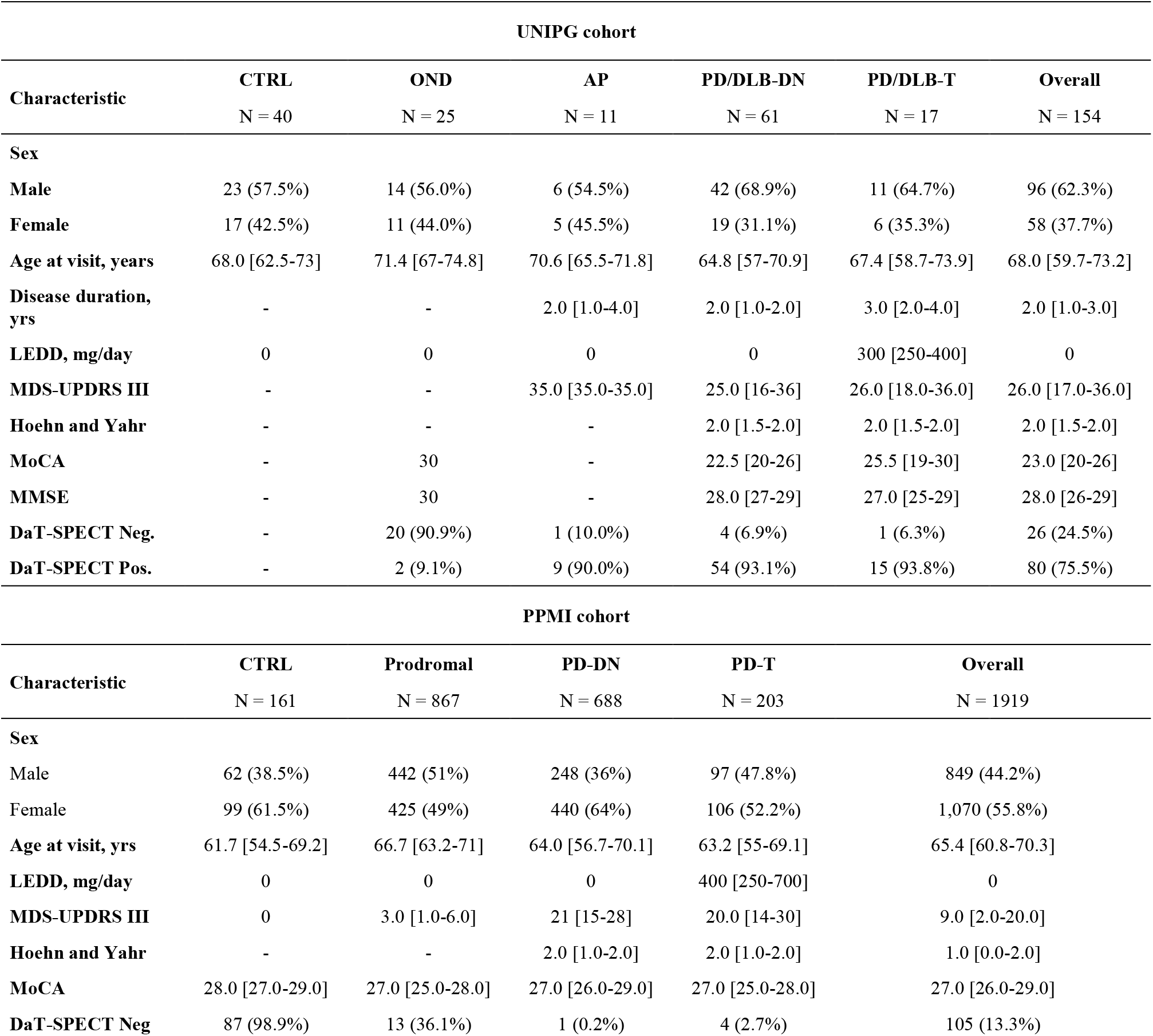
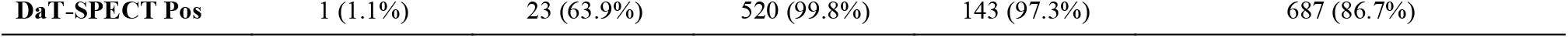
Characteristics of the UNIPG and PPMI cohort. Data are presented as median [first quartile, third quartile] for continuous variables and n (%) for categorical variables. Percentages were calculated among participants with available data for each variable. Dashes indicate unavailable or non-applicable data. AP, atypical parkinsonian syndromes; CTRL, healthy controls; DaT-SPECT, dopamine transporter single-photon emission computed tomography; DLB, dementia with Lewy bodies; LEDD, levodopa equivalent daily dose; MDS-UPDRS III, Movement Disorder Society Unified Parkinson’s Disease Rating Scale part III; MMSE, Mini-Mental State Examination; MoCA, Montreal Cognitive Assessment; OND, other neurological diseases; PD, Parkinson’s disease; PPMI, Parkinson’s Progression Markers Initiative.

| UNIPG cohort |  |  |  |  |  |  |
| --- | --- | --- | --- | --- | --- | --- |
| Characteristic | CTRL<br>N = 40 | OND<br>N = 25 | AP<br>N = 11 | PD/DLB-DN<br>N = 61 | PD/DLB-T<br>N = 17 | Overall<br>N = 154 |
| Sex |  |  |  |  |  |  |
| Male | 23 (57.5%) | 14 (56.0%) | 6 (54.5%) | 42 (68.9%) | 11 (64.7%) | 96 (62.3%) |
| Female | 17 (42.5%) | 11 (44.0%) | 5 (45.5%) | 19 (31.1%) | 6 (35.3%) | 58 (37.7%) |
| Age at visit, years | 68.0 [62.5-73] | 71.4 [67-74.8] | 70.6 [65.5-71.8] | 64.8 [57-70.9] | 67.4 [58.7-73.9] | 68.0 [59.7-73.2] |
| Disease duration, yrs | - | - | 2.0 [1.0-4.0] | 2.0 [1.0-2.0] | 3.0 [2.0-4.0] | 2.0 [1.0-3.0] |
| LEDD, mg/day | 0 | 0 | 0 | 0 | 300 [250-400] | 0 |
| MDS-UPDRS III | - | - | 35.0 [35.0-35.0] | 25.0 [16-36] | 26.0 [18.0-36.0] | 26.0 [17.0-36.0] |
| Hoehn and Yahr | - | - | - | 2.0 [1.5-2.0] | 2.0 [1.5-2.0] | 2.0 [1.5-2.0] |
| MoCA | - | 30 | - | 22.5 [20-26] | 25.5 [19-30] | 23.0 [20-26] |
| MMSE | - | 30 | - | 28.0 [27-29] | 27.0 [25-29] | 28.0 [26-29] |
| DaT-SPECT Neg. | - | 20 (90.9%) | 1 (10.0%) | 4 (6.9%) | 1 (6.3%) | 26 (24.5%) |
| DaT-SPECT Pos. | - | 2 (9.1%) | 9 (90.0%) | 54 (93.1%) | 15 (93.8%) | 80 (75.5%) |
| PPMI cohort |  |  |  |  |  |  |
| Characteristic | CTRL<br>N = 161 | Prodromal<br>N = 867 | PD-DN<br>N = 688 | PD-T<br>N = 203 | Overall<br>N = 1919 |  |
| Sex |  |  |  |  |  |  |
| Male | 62 (38.5%) | 442 (51%) | 248 (36%) | 97 (47.8%) | 849 (44.2%) |  |
| Female | 99 (61.5%) | 425 (49%) | 440 (64%) | 106 (52.2%) | 1,070 (55.8%) |  |
| Age at visit, yrs | 61.7 [54.5-69.2] | 66.7 [63.2-71] | 64.0 [56.7-70.1] | 63.2 [55-69.1] | 65.4 [60.8-70.3] |  |
| LEDD, mg/day | 0 | 0 | 0 | 400 [250-700] | 0 |  |
| MDS-UPDRS III | 0 | 3.0 [1.0-6.0] | 21 [15-28] | 20.0 [14-30] | 9.0 [2.0-20.0] |  |
| Hoehn and Yahr | - | - | 2.0 [1.0-2.0] | 2.0 [1.0-2.0] | 1.0 [0.0-2.0] |  |
| MoCA | 28.0 [27.0-29.0] | 27.0 [25.0-28.0] | 27.0 [26.0-29.0] | 27.0 [25.0-28.0] | 27.0 [26.0-29.0] |  |
| DaT-SPECT Neg | 87 (98.9%) | 13 (36.1%) | 1 (0.2%) | 4 (2.7%) | 105 (13.3%) |  |

**Table 1. Characteristics of the UNIPG and PPMI cohort.**
|  |  |  |  |  |  |
| --- | --- | --- | --- | --- | --- |
| **DaT-SPECT Pos** | 1 (1.1%) | 23 (63.9%) | 520 (99.8%) | 143 (97.3%) | 687 (86.7%) |

Clinical and imaging characteristics followed the expected patterns across diagnostic groups. In PPMI, age and sex distributions differed across groups (both p < 0.001); motor impairment was greater in PD than in prodromal participants, while differences in median cognitive scores were small. DaT-SPECT positivity was nearly universal in PD, intermediate in prodromal participants, and rare in controls (p < 0.001). The composition of the prodromal group is reported in **Supplementary Table 3**. In UNIPG, age and sex distributions did not differ significantly across groups (p = 0.069 and p = 0.689, respectively), and motor and cognitive scores were similar between drug-naïve and treated patients. DaT-SPECT scans were predominantly positive in PD/DLB and AP and negative in OND (p < 0.001).

### CSF DDC is the protein most associated with DaT-SPECT imaging in the PPMI cohort

We used a linear regression model, corrected for age, sex and dopaminergic treatment, to determine the proteins mostly associated with DaT-SPECT SBR (n=1605, control, prodromal and PD subjects). Among the 129 proteins assessed with the CNS NULISA panel, DDC showed the highest degree of association with SBR for the total striatum (β = −0.132, adj. *p* < 0.001) (**Supplementary Figure 1**). When caudate and putamen were analyzed separately, DDC was still the top-ranking protein with a negative β value of –0.195 (adj. *p* <0.001) in the putamen, while the association was weaker in the caudate, where DDC was ranked fourth with a β value of –0.060 (adj. *p* <0.001).

**Figure 1.**
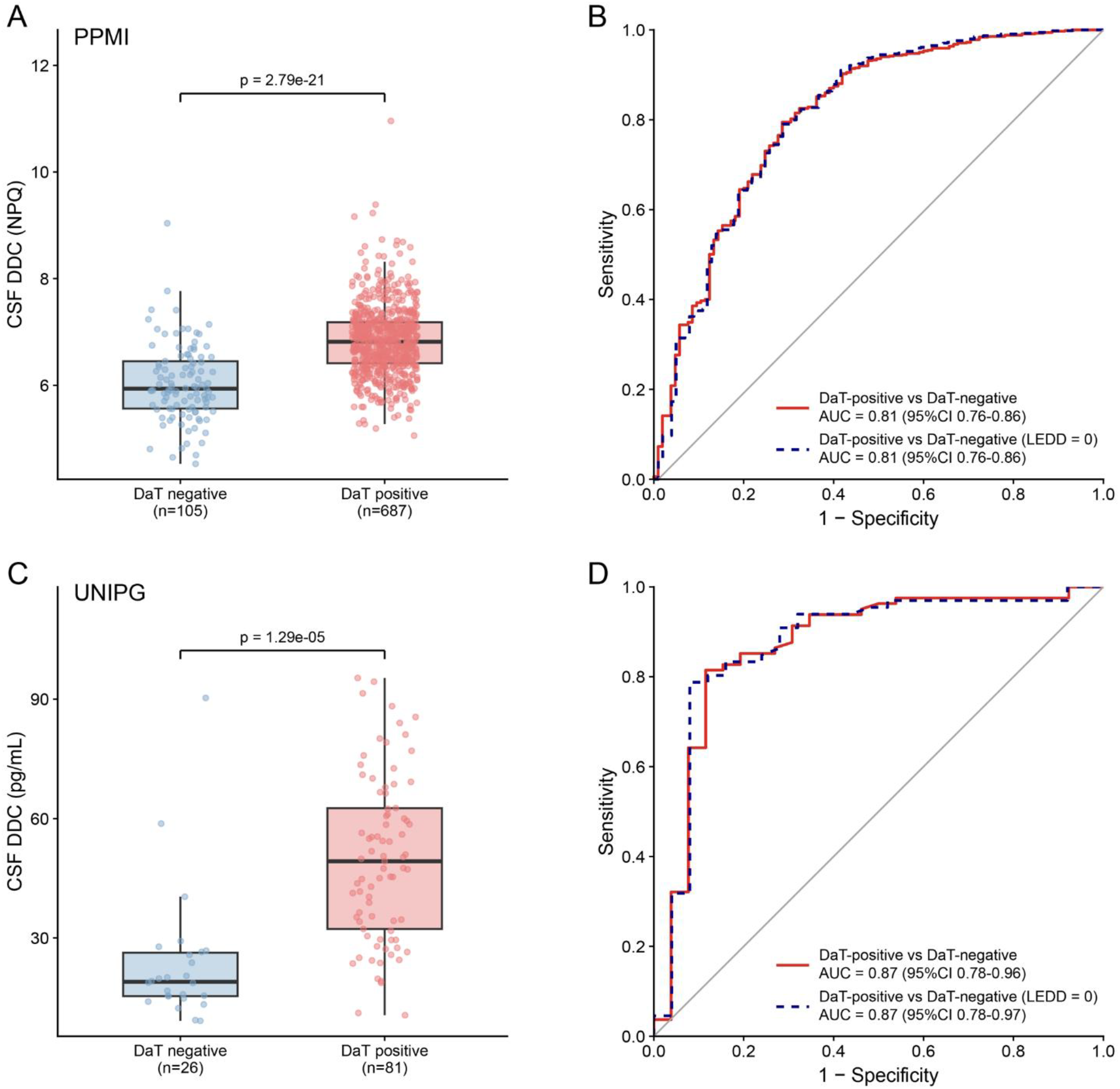
CSF DDC levels according to qualitative DaT-SPECT results in the PPMI and UNIPG cohorts. **(A)** CSF DDC levels (NPQ) in DaT-SPECT− and DaT-SPECT+ participants from the PPMI cohort. Horizontal lines indicate medians, boxes the IQR, whiskers 1.5 × IQR, and dots individual participants. The reported P value refers to the CSF DDC coefficient in a logistic regression model adjusted for age and LEDD. **(B)** ROC curves showing the performance of CSF DDC in discriminating DaT-SPECT+ from DaT-SPECT− participants in the PPMI cohort. The solid red line represents the entire cohort, whereas the dashed blue line was obtained by excluding PD-T. AUC values are reported with 95% CIs. **(C)** CSF DDC concentrations measured using a fully automated quantitative immunoassay in DaT-SPECT negative and DaT-SPECT positive individuals from the UNIPG cohort. Box plots are displayed as in panel A. The reported P value refers to the CSF DDC coefficient in a logistic regression model adjusted for age and LEDD. **(D)** ROC curves showing the performance of CSF DDC in discriminating DaT-SPECT positive from DaT-SPECT negative individuals in the UNIPG cohort. The solid red line represents the entire cohort, whereas the dashed blue was obtained by excluding PD/DLB-T. Abbreviations: AUC, area under the curve; CI, confidence interval; CSF, cerebrospinal fluid; DaT-SPECT, dopamine transporter single-photon emission computed tomography; DDC, dopa decarboxylase; DLB, dementia with Lewy bodies; IQR, interquartile range; LEDD, levodopa equivalent daily dose; NPQ, normalized protein quantity; PD, Parkinson’s disease; PD-T, treated Parkinson’s disease; PD/DLB-T, treated Parkinson’s disease or dementia with Lewy bodies; PPMI, Parkinson’s Progression Markers Initiative; ROC, receiver operating characteristic.

### CSF DDC discriminates between DaT-SPECT positive vs. DaT-SPECT negative individuals in PPMI and UNIPG cohorts

Median CSF DDC levels were higher in DaT-SPECT-positive than in DaT-SPECT-negative participants in both PPMI (6.82 versus 5.94 NPQ) and UNIPG (47.34 versus 19.46 pg/mL). Higher DDC was associated with DaT-SPECT positivity after adjustment for age and LEDD in both cohorts (both *p* < 0.001; **Figure 1A,C**).

DDC discriminated positive from negative scans in PPMI (AUC 0.811, 95% CI 0.762–0.859) and UNIPG (AUC 0.870, 95% CI 0.777–0.963). Performance remained similar after excluding treated PD patients in PPMI (AUC 0.810, 95% CI 0.758–0.861) and treated PD/DLB patients in UNIPG (AUC 0.868, 95% CI 0.772–0.965; Fig. 1B,D). In UNIPG, the Youden-derived cut-off of 29.34 pg/mL yielded 81.5% sensitivity and 88.5% specificity.

### CSF DDC levels are negatively associated with DaT-SPECT SBR in PPMI and UNIPG cohorts

CSF DDC correlated inversely with mean striatal SBR in PPMI PD + CTRL participants (ρ = −0.377), drug-naïve PD (ρ = −0.162), and treated PD (ρ = −0.360; all *p* < 0.001), with the strongest regional correlations observed for putamen SBR. In UNIPG, inverse correlations were observed in the overall cohort (ρ = −0.466, *p* < 0.001) and drug-naïve PD patients (ρ = −0.392, *p* = 0.0028), with similar findings for putamen and caudate SBR. Among treated PD/DLB patients (n = 16), only the caudate correlation reached significance (ρ = −0.552, *p* = 0.0408; Figure 2).

**Figure 2.**
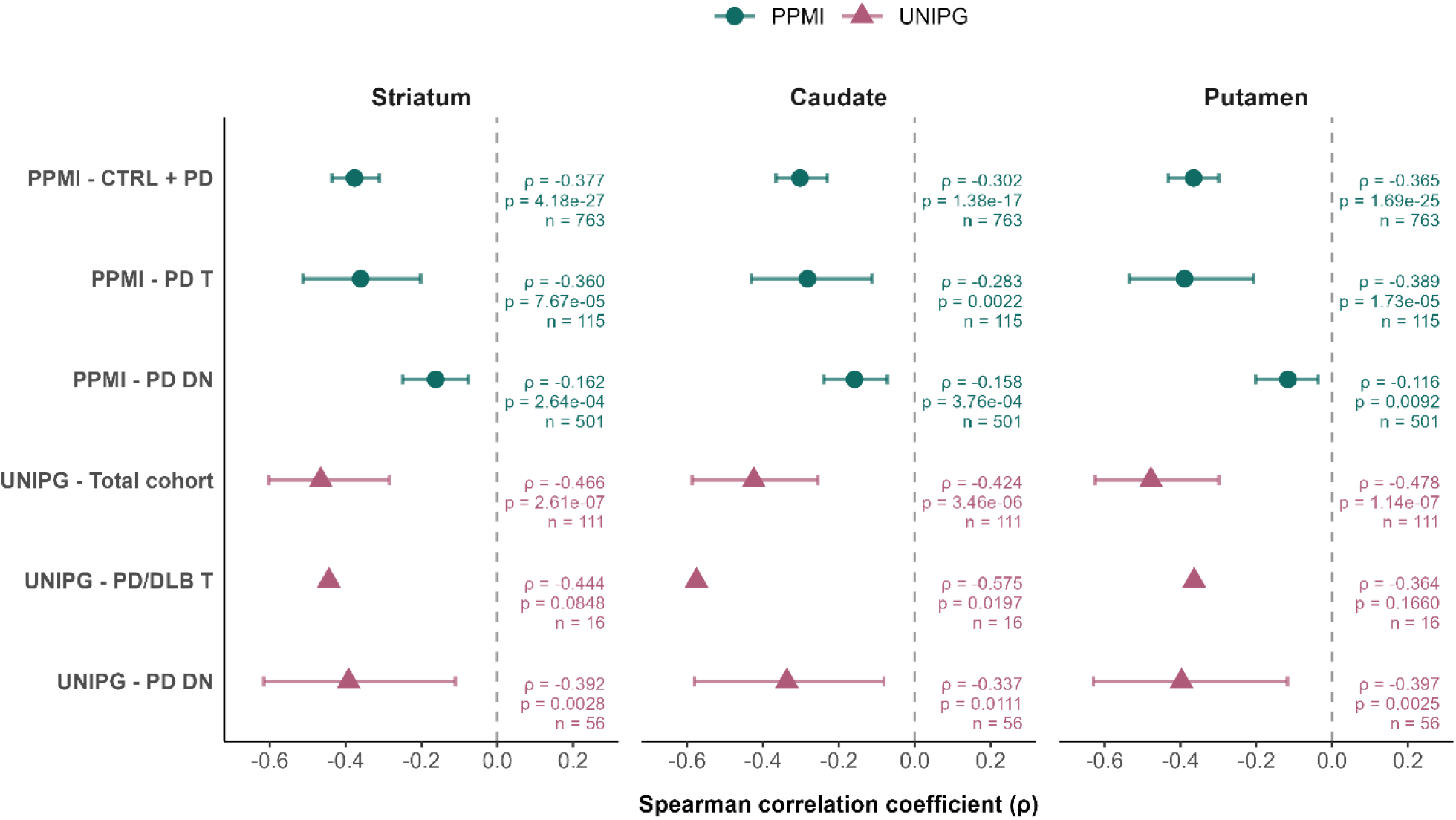
Association between CSF DDC and DaT-SPECT SBR in the PPMI and UNIPG cohorts. Forest plots showing the association between CSF DDC and mean striatal, caudate, and putaminal SBR in the PPMI CTRL + PD, PD T, and PD DN groups, and in the UNIPG total, PD/DLB T, and PD/DLB DN groups. Points represent Spearman’s ρ and horizontal bars indicate 95% bootstrap CIs. Circles denote PPMI estimates and triangles denote UNIPG estimates. Corresponding Spearman’s ρ, two-sided *P* values, and sample sizes are reported for each analysis. **Abbreviations:** CI, confidence interval; CSF, cerebrospinal fluid; CTRL, controls; DaT, dopamine transporter; DDC, dopa decarboxylase; DLB, dementia with Lewy bodies; PD, Parkinson’s disease; PD DN, drug-naïve Parkinson’s disease; PD T, treated Parkinson’s disease; PPMI, Parkinson’s Progression Markers Initiative; SBR, specific binding ratio; UNIPG, University of Perugia.

### CSF DDC is increased across parkinsonian disorders in both PPMI and UNIPG cohorts

In PPMI, CSF DDC levels were higher in prodromal participants and both PD groups than in controls, and in both PD groups than in prodromal participants (all FDR-adjusted *p* < 0.001). Treated PD patients showed a modest increase compared with drug-naïve patients (FDR-adjusted *p* = 0.046; Figure 3A). DDC discriminated PD from controls (AUC 0.855, 95% CI 0.823–0.886), with similar performance in drug-naïve patients (AUC 0.856, 95% CI 0.823–0.889; Figure 3B).

**Figure 3.**
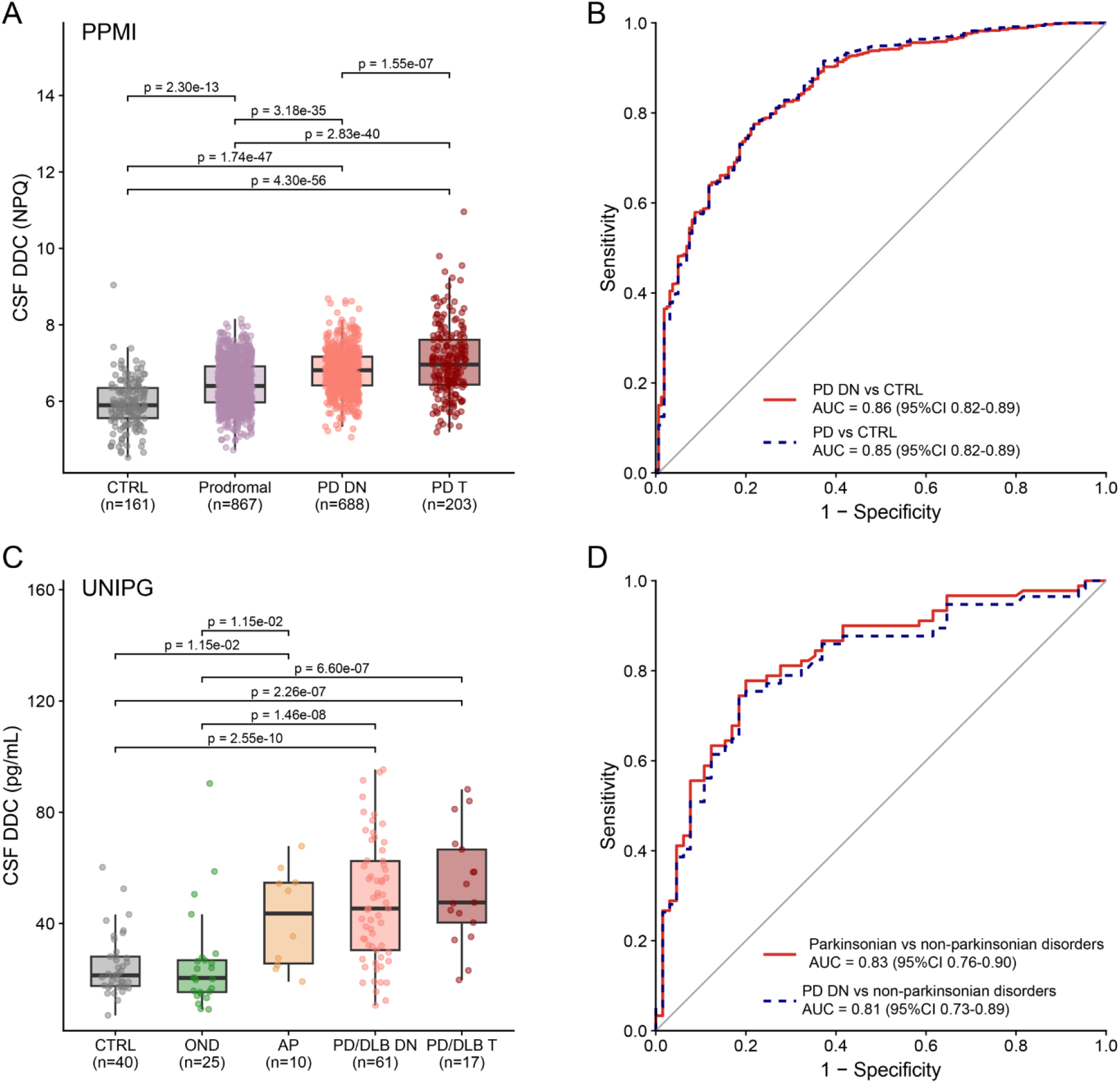
CSF DDC across diagnostic and treatment groups in the PPMI and UNIPG cohorts. (A) CSF DDC, expressed as NPQ, across CTRL, prodromal, PD DN, and PD T groups in PPMI. Horizontal lines indicate medians, boxes represent the IQR, whiskers extend to the most extreme observations within 1.5 × IQR of the box limits, and dots represent individual participants. Pairwise comparisons of estimated marginal means were performed using linear regression models adjusted for age, with Benjamini–Hochberg FDR correction. Only significant comparisons are displayed, up to a maximum of eight per panel. (B) ROC curves for the discrimination of PD from CTRL (solid red line) and PD DN from CTRL (dashed blue line) in PPMI. (C) CSF DDC concentrations, expressed as pg/mL and measured using a fully automated quantitative immunoassay, across CTRL, OND, AP, PD/DLB DN, and PD/DLB T groups in the independent UNIPG cohort. Box plots and statistical comparisons are presented as in panel A. (D) ROC curves for the discrimination of parkinsonian disorders, including AP, PD, and DLB, from non-parkinsonian conditions, including CTRL and OND, in UNIPG. The solid red line represents all parkinsonian disorders irrespective of treatment status, whereas the dashed blue line represents drug-naïve PD patients versus non-parkinsonian conditions. In panels B and D, AUCs are reported with two-sided 95% CIs calculated using the DeLong method. Abbreviations: AP, atypical parkinsonian syndromes; AUC, area under the curve; CI, confidence interval; CSF, cerebrospinal fluid; CTRL, controls; DDC, dopa decarboxylase; DLB, dementia with Lewy bodies; FDR, false discovery rate; IQR, interquartile range; NPQ, normalized protein quantity; OND, other neurological diseases; PD, Parkinson’s disease; PD DN, drug-naïve Parkinson’s disease; PD T, treated Parkinson’s disease; PD/DLB DN, drug-naïve Parkinson’s disease or dementia with Lewy bodies; PD/DLB T, treated Parkinson’s disease or dementia with Lewy bodies; PPMI, Parkinson’s Progression Markers Initiative; ROC, receiver operating characteristic; UNIPG, University of Perugia.

In UNIPG, DDC levels were higher in both PD/DLB groups than in CTRL and OND (all FDR-adjusted *p* < 0.001). AP patients also had higher levels than CTRL and OND (both FDR-adjusted *p* = 0.016), whereas differences between AP and either PD/DLB group, or between drug-naïve and treated PD/DLB patients, were not significant (Figure 3C). DDC discriminated parkinsonian disorders from CTRL and OND (AUC 0.830, 95% CI 0.764–0.896), with similar performance when cases were restricted to drug-naïve PD patients (AUC 0.811, 95% CI 0.732–0.890; Figure 3D).

### CSF DDC, like DaT-SPECT, predicts progression to motor symptoms in prodromal PD

Among 627 prodromal PPMI participants with baseline CSF DDC and DaT-SPECT measurements, 100 (16.0%) reached an MDS-UPDRS III score ≥10 during follow-up of up to six years.

Higher baseline DDC was associated with motor progression both above versus below the median (adjusted HR 1.85, 95% CI 1.21–2.83; *p* = 0.004; log-rank *p* < 0.001) and per SD increase (adjusted HR 1.41, 95% CI 1.15–1.73; *p* = 0.001; Figure 4A). Higher putamen SBR was associated with lower progression hazard in both median-dichotomized (adjusted HR 0.63, 95% CI 0.41–0.96; *p* = 0.030) and continuous analyses (HR per SD 0.65, 95% CI 0.53–0.80; *p* < 0.001; Figure 4B). Total striatal SBR was associated with progression only in the continuous analysis (HR per SD 0.71, 95% CI 0.57–0.88; *p* = 0.001), whereas the median-based comparison was not significant (HR 0.68, 95% CI 0.44–1.04; *p* = 0.074; Figure 4D). For caudate SBR, the log-rank test was significant (*p* = 0.012), but the adjusted median-based association was not (HR 0.91, 95% CI 0.58–1.43; *p* = 0.681; Figure 4C).

**Figure 4.**
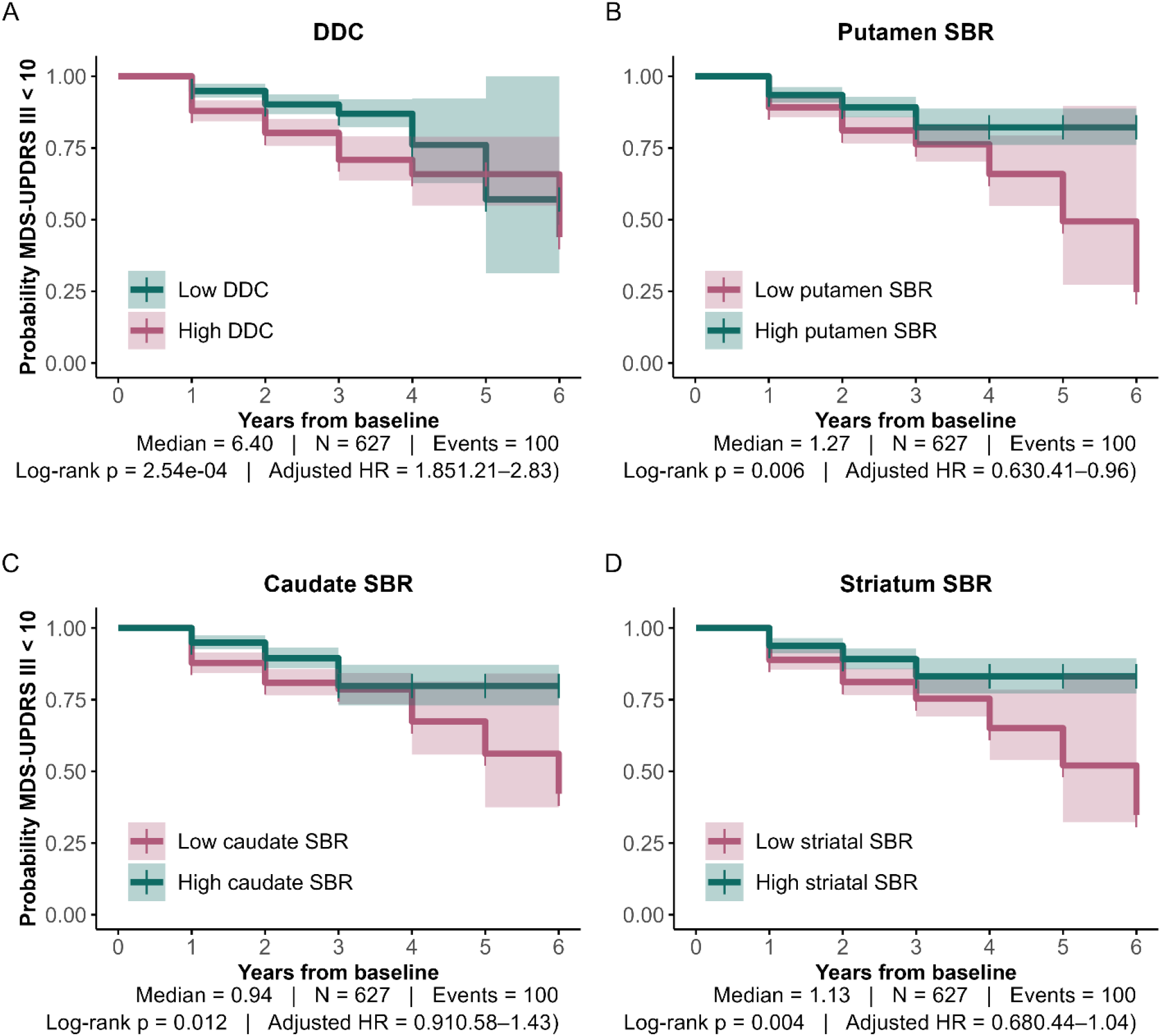
Prognostic value of CSF DDC and DaT-SPECT SBR for motor progression in prodromal PD. Kaplan– Meier curves showing the probability of remaining below an MDS-UPDRS III score of 10 in prodromal participants from the PPMI cohort. Participants were dichotomized according to the median baseline value for CSF DDC (A), putamen SBR (B), caudate SBR (C), and total striatal SBR (D). Shaded areas represent 95% CIs. HRs were estimated using Cox proportional hazards models adjusted for age, sex, prodromal subgroup, and baseline MDS-UPDRS III score. *P* values were calculated using two-sided log-rank tests. Higher CSF DDC levels and lower striatal DaT-SPECT uptake were associated with an increased risk of motor progression. Abbreviations: CI, confidence interval; CSF, cerebrospinal fluid; DaT, dopamine transporter; DDC, dopa decarboxylase; HR, hazard ratio; MDS-UPDRS III, Movement Disorder Society– Unified Parkinson’s Disease Rating Scale part III; PD, Parkinson’s disease; PPMI, Parkinson’s Progression Markers Initiative; SBR, specific binding ratio.

### CSF DDC is associated with motor and cognitive scores in drug-naïve PD patients in both PPMI and UNIPG cohorts

In the PPMI cohort, higher CSF DDC levels correlated with greater motor impairment, as assessed by MDS-UPDRS III, in both drug-naïve (ρ = 0.196, *p* < 0.001) and treated patients (ρ = 0.213, *p* = 0.002). Similar findings were observed in the independent UNIPG cohort in drug-naïve patients (ρ = 0.370, *p* = 0.009), whereas the corresponding correlation in treated patients did not reach statistical significance, likely due to the limited sample size. By contrast, associations with global cognition were restricted to drug-naïve patients. Higher CSF DDC levels were associated with lower MoCA scores in both the PPMI (ρ = −0.093, *p* = 0.014) and UNIPG cohorts (ρ = −0.427, *p* = 0.004), whereas no significant correlations were observed among treated patients in either cohort (**Supplementary Figure 2**).

## Discussion

In the present study, we provide evidence that CSF DDC is a promising fluid biomarker of nigrostriatal dopaminergic dysfunction. Using affinity proteomic data from the PPMI, DDC was the protein most strongly associated with qualitative and quantitative DaT-SPECT measures. We replicated these findings in an independent real-world cohort using a fully automated quantitative chemiluminescent immunoassay. Despite substantial differences between the two cohorts, CSF DDC consistently identified individuals with dopaminergic dysfunction, showing an inverse correlation with SBR, and was also increased in all parkinsonian disorders. In the PPMI cohort, CSF DDC was also increased in prodromal PD and predicted the onset of motor symptoms.

Previous evidence on the relationship between CSF DDC and DaT-SPECT qualitative outcome has been inconclusive. A recent study using ultrasensitive immunoassays found no significant differences in CSF DDC between individuals with normal and abnormal DaT-SPECT,^5^ although the analysis was restricted to a small subset of DLB patients without available information on dopaminergic treatment. Conversely, another study including PPMI patients, reported that DDC can discriminate PD from people with scans without evidence of dopaminergic deficit (SWEDD) and healthy controls^7^. Similarly, studies examining quantitative DaT-SPECT measures yielded conflicting results^6,7^, possibly reflecting differences in diagnostic composition, analytical methods for DDC measurements and DaT-SPECT assessment.

Our findings show, in two independent cohorts, that CSF DDC is associated with both qualitative and quantitative measures of dopaminergic dysfunction, discriminating DaT-SPECT-positive from DaT-SPECT-negative individuals and inversely correlating with SBR.

In both cohorts, this association was mostly driven by putamen SBR, potentially reflecting the typical topography of nigrostriatal degeneration in PD, which preferentially affects dopaminergic projections to the putamen, sparing the caudate nucleus in earlier disease stages^14,15^. Together, these findings suggest that DDC might be able to capture both the presence and severity of nigrostriatal dopaminergic dysfunction.

The potential effect of dopaminergic treatment is particularly relevant when considering DDC as a biomarker. During L-DOPA treatment, peripheral DDC is inhibited using carbidopa or benserazide to reduce peripheral L-DOPA decarboxylation and increase its transport across the blood-brain barrier^16^. Previous studies suggested that blood DDC concentrations and/or activity is influenced by dopaminergic therapy,^5^ possibly as consequence of the inhibition of peripheral DDC,^17,18^ limiting its usefulness as a peripheral marker of dopaminergic dysfunction^19,20^. In contrast, in our study the ability of CSF DDC to discriminate DaT-SPECT-positive from DaT-SPECT-negative individuals and parkinsonian from non-parkinsonian conditions was substantially independent of dopaminergic treatment.

Another important finding of the present study concerns CSF DDC levels during the prodromal phase of PD. In prodromal PD, DDC levels were intermediate between those observed in healthy controls and patients with established PD and predicted the onset of motor symptoms with performance comparable to or better than DaT-SPECT measures. These results are consistent with previous evidence of increased CSF DDC in the preclinical Lewy body diseases, i.e., α-syn-SAA positive individuals without any symptoms, where higher DDC levels were associated with an increased risk of progression to symptomatic disease.^4^ In this context, increased CSF DCC may represent an early biochemical signature of nigrostriatal dopaminergic dysfunction caused by α-synuclein pathology, possibly preceding the onset of clinical symptoms. Further supporting the biological relevance of CSF DDC as marker of nigrostriatal dopaminergic dysfunction, higher DDC was consistently associated with more severe motor impairment in drug-naïve PD patients in both cohorts. This is particularly relevant as the cardinal motor manifestations of PD largely arise from progressive dysfunction of nigrostriatal dopaminergic pathway^21^. In drug-naïve patients of both cohorts, higher CSF DDC levels were associated with poorer global cognitive performance as assessed by MoCA, which is particularly sensitive to executive deficits that are prominent in PD and partly related to the dysfunction of dopaminergic fronto-striatal circuits^22^.

Within proposed biological frameworks for PD, CSF may enable the simultaneous detection of distinct pathogenetic processes associated with the disease. While CSF α-syn-SAA is used to assess synucleinopathy (S), our findings support the role of CSF DDC as a potential biochemical counterpart of DaT-SPECT for defining dopaminergic impairment (D).

Further validation is required before CSF DDC can be used in routine clinical practice. So far, evidence supporting DDC as a PD biomarker has been mostly generated using proteomic platforms providing relative measures of protein abundance; this approach is valuable for biomarker discovery but not suitable to routine clinical implementation^23,24,25,26,27^. A major strength of this study is the replication of PPMI proteomic findings in an independent real-world cohort using a quantitative, fully automated chemiluminescent immunoassay. This provides a basis for future studies aimed at assessing inter-laboratory performance and establishing reference ranges across clinical populations. Several limitations should nevertheless be considered. The PPMI and UNIPG cohorts differed in recruitment setting, diagnostic composition, sample size, and biomarker methodology. Although this heterogeneity strengthens the generalizability of the replicated associations, absolute DDC values could not be directly compared between cohorts. Besides dopaminergic replacement therapy, the potential influence of psychoactive medications was not comprehensively assessed. This may be relevant for antipsychotic drugs with anti-dopaminergic properties and for selective serotonin reuptake inhibitors, which may interfere with DaT-SPECT measurement^28^ and may theoretically influence DDC in CSF, since this enzyme is also involved in serotonin synthesis^3^.

Overall, our findings indicate that CSF DDC provides a quantitative biochemical measure of nigrostriatal dopaminergic dysfunction that closely parallels DaT-SPECT across different clinical settings and analytical approaches. Its increase in prodromal disease, association with subsequent motor progression, and measurement using a fully automated quantitative assay further support its translational potential. Together with α-syn-SAA, CSF DDC may contribute to a scalable fluid biomarker framework for the biological characterization and staging of PD.

## Data availability

PPMI data used in this study are available through the Parkinson’s Progression Markers Initiative database (www.ppmi-info.org/access-data-specimens/download-data; RRID: SCR_006431). DaT-SPECT data for prodromal participants were obtained from PPMI upon request after approval by the PPMI Data Access Committee. Data from the UNIPG cohort are available from the corresponding authors upon reasonable request, subject to applicable ethical and data protection regulations.

## Acknowledgements

Data used in the preparation of this article were obtained on 2026-06-15 from the Parkinson’s Progression Markers Initiative (PPMI) database (www.ppmi-info.org/access-data-specimens/download-data), RRID:SCR_006431. For up-to-date information on the study, visit www.ppmi-info.org. PPMI – a public-private partnership – is funded by the Michael J. Fox Foundation for Parkinson’s Research and funding partners, including AbbVie, Alamar Biosciences, Aligning Science Across Parkinson’s (ASAP), Arrowhead Pharma, Arvinas, AskBio, BIAL, BioArctic, Biohaven, BlueRock Therapeutics, Bristol Myers Squibb, Calico Labs, Capsida Biotherapeutics, Critical Path Institute, DaCapo Brainscience, Denali, Edmond J. Safra Foundation, Eli Lilly, Gain Therapeutics, GE Healthcare, Genentech, GSK, Insitro, Johnson & Johnson Innovative Medicine, Lundbeck, Merck, Neumora, Neuron23, Novartis, Olink, Regeneron, Roche, Sanofi, Tenvie, UCB, Vanqua Bio, Voyager Therapeutics, The Weston Family Foundation.

We would like to acknowledge the support of the Miriade consortium, in particular Prof. Charlotte Teunissen and Dr. Katharina Bolsewig for their support during the proof-of-concept DDC assay formats.

