## Supplementary for "Cerebrospinal fluid dopa-decarboxylase is a biomarker of dopaminergic dysfunction"

### Supplementary materials

#### Supplementary Methods

##### PPMI dataset construction

PPMI data were obtained on June 15, 2026. DaT-SPECT data for prodromal participants were released upon request after approval by the PPMI Data Access Committee. Baseline CSF proteomic measurements were obtained using the CNS 120 NULISA® panel (Alamar Biosciences, Fremont, CA, USA; Project 282: High-Plex Profiling of Neuro-Related Proteins in Human CSF with NULISA [CNS Panel]). When multiple DaT-SPECT scans were available, the scan closest to CSF collection and within 18 months was selected. Availability of proteomic and DaT-SPECT data across diagnostic groups is reported in Supplementary Table 1. Demographic and clinical variables included age, sex, disease duration, dopaminergic treatment status, total levodopa equivalent daily dose (LEDD), MDS-UPDRS Part III, Hoehn and Yahr stage and Montreal Cognitive Assessment (MoCA).

##### PPMI DaT-SPECT imaging

DaT-SPECT images were acquired at participating PPMI centres according to a standardized protocol and centrally processed by the PPMI imaging core laboratory. Visual assessments classified scans as positive or negative for evidence of a presynaptic dopamine transporter deficit. Bilateral caudate and putaminal SBR values were calculated by averaging left- and right-sided measurements. The whole-striatum SBR provided by the imaging core laboratory was used as the global measure of striatal dopamine transporter binding.

##### UNIPG cohort: clinical data and DaT-SPECT

Demographic variables (age and sex) were available for the whole cohort. For patients with parkinsonian syndromes, disease duration, dopaminergic treatment, LEDD, MDS-UPDRS Part III, Hoehn and Yahr stage, MoCA and Mini-Mental State Examination (MMSE) scores were also collected. The final diagnosis of OND was based on clinical and imaging findings.
DaT-SPECT scans were acquired 3–4 h after intravenous administration of $^{123}$I-ioflupane (DaTSCAN). Scans were visually classified by an expert reader as positive or negative for presynaptic dopaminergic dysfunction. Quantitative analysis was performed with Basal Ganglia Matching Tools V2, based on the automated three-dimensional BasGan method. SBR values were obtained separately for the left and right caudate nuclei and putamina using an occipital background reference and partial volume effect correction. Bilateral caudate and putamen SBR values were calculated by averaging

the corresponding left- and right-sided measurements; mean striatal SBR was calculated as the average of bilateral caudate and putamen SBR values.

##### Lumipulse® G DDC assay: analytical qualification

The assay is calibrated with recombinant full-length DDC protein, with a calibration range of 1.5–600 pg/mL. The functional measurement range is 1.5–250 pg/mL and the lower functional limit of quantification (LLOQ) is 0.5 pg/mL. Precision was assessed on 5 samples (2 replicates over 5 runs), with mean intra- and inter-assay variation of 3.8% and 4.3%, respectively. Parallelism was assessed using CSF samples from six individuals at dilution factors from 1 to 6. Dilution-corrected concentrations were consistent across the series, indicating parallel behaviour between endogenous CSF DDC and the recombinant calibrator and no matrix interference. Spike recovery was evaluated by adding recombinant DDC to human CSF at 9, 30 and 75 pg/mL, and all spikes met the predefined acceptance criteria (90–110%).

##### Pre-analytical handling and sample analysis (UNIPG)

On the day of analysis, samples were thawed at room temperature for at least 30 minutes and centrifuged (8 min, 10,000 × g, 4 °C). Seventy microlitres of CSF were transferred into sample cups and diluted 1:3 with assay buffer according to the assay protocol. All samples were analysed in single measurements within one analytical run. Kit quality control samples were included and met predefined acceptance criteria. Measurements were performed blind to clinical data.

##### Statistical analysis: additional details

In the proteome-wide analyses, standardized regression coefficients were obtained after z-standardization of both protein and imaging variables. In logistic regression models, qualitative DaT-SPECT status was the dependent variable and CSF DDC the predictor. Time-to-event analyses included prodromal participants with available imaging and proteomic measurements at the baseline visit. Follow-up was administratively censored at 6 years, and participants who did not experience the endpoint were censored at their last available visit. For Kaplan–Meier analyses, participants were divided into two groups using the median of CSF DDC, putamen, caudate and total striatal SBR, separately. In Cox models, continuous biomarkers were standardized per 1 SD increase.

#### Supplementary Tables

**Supplementary Table 1.** Availability of CSF proteomic and DaT-SPECT data in the PPMI cohort.

| | PD | Prodromal PD | CTRL | Total |
|---|---|---|---|---|
| CSF NULISA proteomics (baseline), n | 891 | 867 | 161 | 1,919 |
| Quantitative DaT-SPECT (SBR), n (%) | 669 (75.1) | 843 (97.2) | 94 (58.4) | 1,606 (83.7) |
| Qualitative DaT-SPECT (visual read), n (%) | 668 (75.0) | 36 (4.2) | 88 (54.7) | 792 (41.3) |

Percentages refer to participants with baseline CSF NULISA proteomic data within each diagnostic group. DaT-SPECT scans were acquired closest to CSF collection and within 18 months. DaT-SPECT data for prodromal participants were obtained upon approval by the PPMI Data Access Committee. CSF, cerebrospinal fluid; CTRL, healthy controls; DaT-SPECT, dopamine transporter single-photon emission computed tomography; PD, Parkinson's disease; PPMI, Parkinson's Progression Markers Initiative; SBR, striatal binding ratio.

**Supplementary Table 2. Composition of the prodromal PD patients from the PPMI cohort.**

| | **N** | **% of Prodromal** |
|---|---|---|
| Hyposmia | 600 | 69.3% |
| RBD | 245 | 28.3% |
| PRKN | 7 | 0.8% |
| LRRK2 | 5 | 0.6% |
| GBA | 5 | 0.6% |
| PRKN + RBD | 2 | 0.2% |
| GBA + RBD | 1 | 0.1% |
| LRRK2 + RBD | 1 | 0.1% |
| Total Prodromal | 866 | 100.0% |

Abbreviations: GBA = glucocerebrosidase gene variant carrier; LRRK2 = leucine-rich repeat kinase 2 gene variant carrier; PRKN = parkin (PRKN) gene mutation; RBD = rapid eye movement sleep behaviour disorder.

**Supplementary Table 3. Composition of the OND group from the UNIPG cohort**

| | N | % of OND |
|---|---|---|
| ET | 7 | 28.0% |
| FTD | 5 | 20.0% |
| AD | 5 | 20.0% |
| Non-AD MCI | 3 | 12.0% |
| Ataxia | 2 | 8.0% |
| NPH | 2 | 8.0% |
| ALS | 1 | 4.0% |
| Total OND | 25 | 100.0% |

Abbreviations: AD = Alzheimer's disease; ALS = amyotrophic lateral sclerosis; ET = essential tremor; FTD = frontotemporal dementia; MCI = mild cognitive impairment; NPH = normal pressure hydrocephalus; OND = other neurological disorders.

#### Supplementary Figures

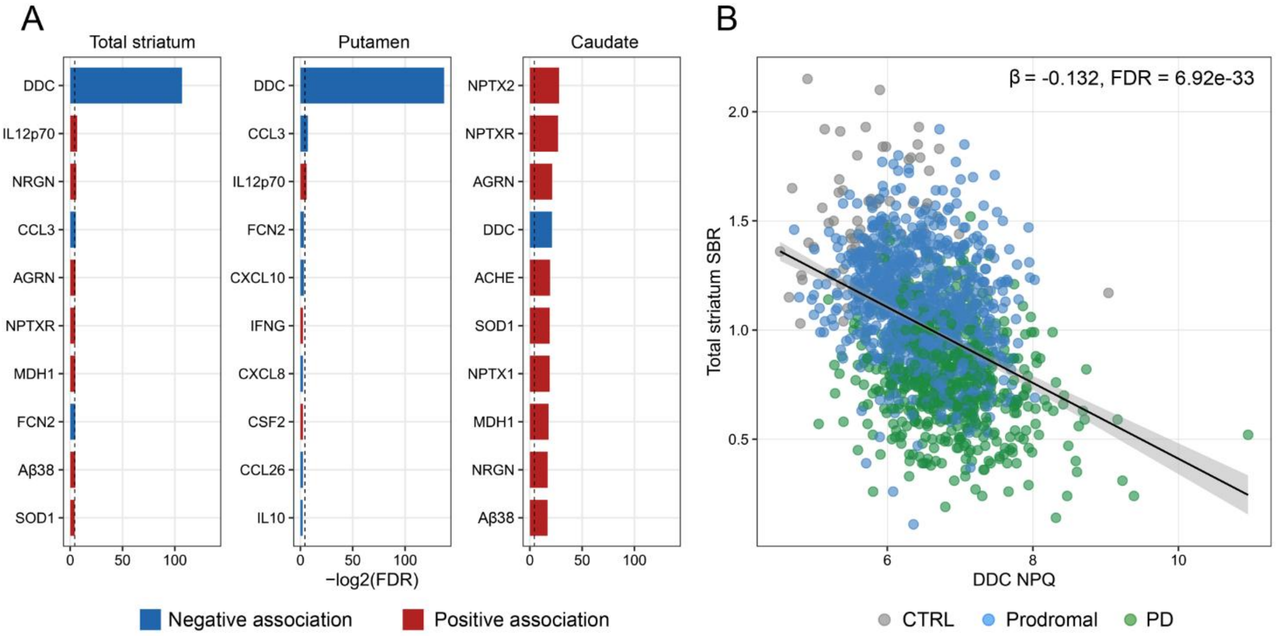


**Supplementary figure 1. Association between CSF NULISA proteins and DaT-SPECT imaging in the PPMI cohort.** A) Ranked associations between baseline CSF NULISA proteins and DAT-SPECT imaging outcomes, including total striatum, putamen, and caudate specific binding ratios. Bars show −log2(FDR) from linear regression models adjusted for age, sex, and total LEDD. Proteins are ordered by FDR within each imaging outcome. Only top ten proteins according to −log2(FDR) are included. Bar colour indicates the direction of association based on the protein beta coefficient. The dashed line indicates FDR = 0.05. B) Association between baseline CSF DDC NPQ levels and total striatal DAT-SPECT specific binding ratio. Points represent individual participants and are coloured by diagnostic group. The regression line shows the unadjusted visual trend, while the reported beta coefficient, standardized beta, and FDR are derived from the linear model adjusted for age, sex, and total LEDD. Abbreviations: CSF, cerebrospinal fluid; CTRL, controls; DaT-SPECT, dopamine transporter single-photon emission computed tomography; DDC, dopa decarboxylase; FDR, false discovery rate; LEDD, levodopa equivalent daily dose; NPQ, normalized protein quantity; PD, Parkinson’s disease.

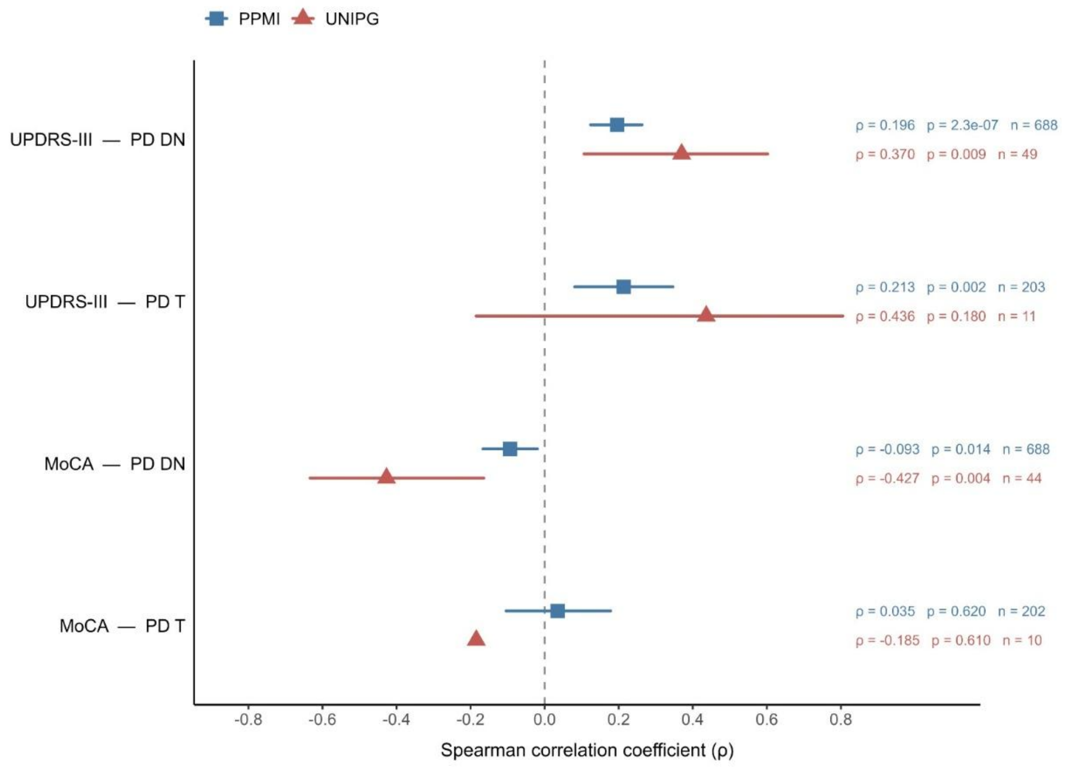


**Supplementary figure 2. Associations among CSF DDC levels and clinical measures (MDS-UPDRS-III and MoCA scores) in PPMI and UNIPG cohorts.** Spearman correlation coefficients (ρ) between CSF dopa decarboxylase (DDC) levels and motor impairment, assessed by the Movement Disorder Society–Unified Parkinson's Disease Rating Scale part III (MDS-UPDRS III), and global cognition, assessed by the Montreal Cognitive Assessment (MoCA), in drug-naïve (PD-DN) and treated (PD-T) Parkinson's disease patients from the Parkinson's Progression Markers Initiative (PPMI; blue squares) and the independent UNIPG cohort (red triangles). Symbols represent Spearman correlation coefficients and horizontal bars indicate 95% confidence intervals. Corresponding ρ values, *P* values and sample sizes are reported alongside each estimate. Across both cohorts, higher CSF DDC levels were consistently associated with greater motor impairment in drug-naïve patients, whereas inverse associations with cognitive performance were observed predominantly in drug-naïve subjects and were not evident in treated patients.
